# The correlation between medial pattern of intracranial arterial calcification and white matter hyperintensities

**DOI:** 10.1101/2023.03.20.23287507

**Authors:** Heng Du, Jianrong Zheng, Xuelong Li, Yanjing Dong, Yajing Cheng, Cong Liu, Jun Hu, Xiangyan Chen

## Abstract

**Background:** Despite previously reported correlations between intracranial arterial calcification (IAC) and white matter hyperintensities (WMH), little is known about the relationship between IAC pattern and WMH. By differentiating intimal and medical IAC, we aimed to investigate the relationship between IAC pattern and WMH.

**Methods:** Consecutive patients with acute ischemic stroke were included. IAC pattern was categorized as intimal or medial on plain brain CT. The number of cerebral arteries involved by IAC for each patient was recorded. IAC severity of each artery was defined as focal or diffuse. On brain MRI, the burden of WMH was graded on a visual rating scale and classified as absent mild, moderate and severe. Multiple logistic regression was performed to examine the relationship between IAC and WMH.

**Results:** Among 265 recruited patients, intimal IAC was detected in 54.7% patients, medial IAC in 48.5% patients and coexistent (intimal and medial) IAC in 52.1% patients. Diffuse IAC was in 27.9% patients, all of which were medial IACs. WMH was found in 75.5% patients, including 105 patients (39.6%) with mild WMH, 69 (26.0%) with moderate WMH and 26 (9.8%) with severe WMH. The presence and severity of medial IAC were correlated with WMH occurrence (*p*<0.001, respectively). Chi-square linear trend suggested the number of arteries involved by medial IAC (*p*<0.001) and the severity of medial IAC (*p*<0.001) were correlated with WMH burden. After adjusting age, hypertension, history of stroke and history of ischemic heart disease, multiple ordinal regression demonstrated a positive correlation between the number of arteries involved by medial IAC (*p*<0.001) and the severity of medial IAC (*p*<0.001) with the overall burden of WMH.

**Conclusions:** Medial IAC was correlated with the burden of WMH. The dose-effect relationship between IAC and WMH suggests the need of further investigations on shared underlying mechanisms of intracranial large artery disease and cerebral small vessel disease.

## 1. Introduction

Intracranial arterial calcification (IAC) is found to be an independent risk factor for stroke^1,2^. Apart from its association with intracranial large artery atherosclerosis^3,4^, IAC may also correlate to cerebral small vessel disease (CSVD)^5,6^, of which white matter hyperintensities (WMH) is one of the common types on neuroimaging^7^. However, current studies about the impact of IAC on WMH are controversial. Despite several reports showing that the presence^8^ and the density^9^ of IAC may be associated WMH, opposite findings also suggested irrelevance between IAC and WMH^10^. IAC mainly involves the intima and the media of intracranial arterial walls^11,12^. Histological and clinical evidence suggested that intimal IAC is associated with atherosclerosis^12^ while medial IAC may have impact on arterial stiffness^13^.

By far, little has been studied on the association between IAC pattern and WMH. We hypothesized that IAC patterns may be correlated with WMH differently. In the present study, we aimed to investigate the association between IAC pattern (intimal IAC and medial IAC) and the presence and burden of WMH.

## 2. Methods and materials

### 2.1 Subjects

This study was approved by the Clinical Research Ethics Committee of the Peking University Shenzhen Hospital and the Ethics Committee of The Hong Kong Polytechnic University. Consecutive patients admitted to the stroke center from November 2019 to August 2021 were recruited retrospectively. The inclusion criteria were: 1) patients above 18 years old who had acute ischemic stroke or transient ischemic attack (TIA); 2) non-contrast brain computed tomography (CT) and magnetic resonance imaging (MRI) were performed within 7 days after symptom onset. The exclusion criteria were as follows: 1) contraindications to either CT or MRI; 2) other causes of white matter lesions, including multiple sclerosis, vasculitis, or connective tissue diseases; 3) patients with critical medical condition, such as head trauma and brain tumor; 4) poor quality of imaging data and incomplete clinical data.

### 2.2 Assessment of intracranial arterial calcification

Intracranial arterial calcification (IAC) was assessed by brain CT. Major intracranial artery segments including the lacerum and cavernous segment (C3-C4) of internal carotid arteries (ICAs), the supraclinoid segment to the communicating segment (C5-C7) of ICAs, M1 segment of middle cerebral arteries (MCAs), proximal and distal part of the intracranial (V4) segment of vertebral arteries (VAs) and the basilar artery (BA) were examined. CT images were assessed by two neurologists with more than 5 years’ experience of neuroimaging (XL.L. and H.D.) who were both blinded to clinical characteristics and MRI images. The presence of IAC was defined as hyperdense foci over 130 Hounsfield units (HU). Based on a previously developed and validated grading scale^14^, IAC was categorized into two patterns: intimal IAC and medial IAC (Figure 1). Circularity (1 for dot, 2 for <90 degrees, 3 for 90-270 degrees and 4 for 270-360 degrees), thickness (1 for thick IAC ≥ 1.5mm and 3 for thin IAC < 1.5mm) and morphology (0 for indistinguishable, 1 for irregular/patchy and 4 for continuous) were assessed and graded. A summed score from 1 to 6 was defined as intimal IAC and 7 to 11 was defined as medial IAC. Based on IAC pattern (intimal or medial), IAC was further evaluated by the number of arteries (NOA) involved by IAC and the severity of IAC in individual artery segment. The NOA involved by IAC in each patient was recorded as the summed number of examined artery segments with IAC. The quartiles (Q1, Q2 and Q3) of the recorded numbers were calculated and then used for grouping in subsequent statistical analysis. The severity of IAC in individual artery segment was defined as focal or diffuse (Figure 1): focal IAC was defined as the trajectory of IAC involving less than 1/3 of the examined artery segment; diffuse IAC was defined as the trajectory of IAC involving more than 1/3 of the examined artery segment.

**Figure 1.**
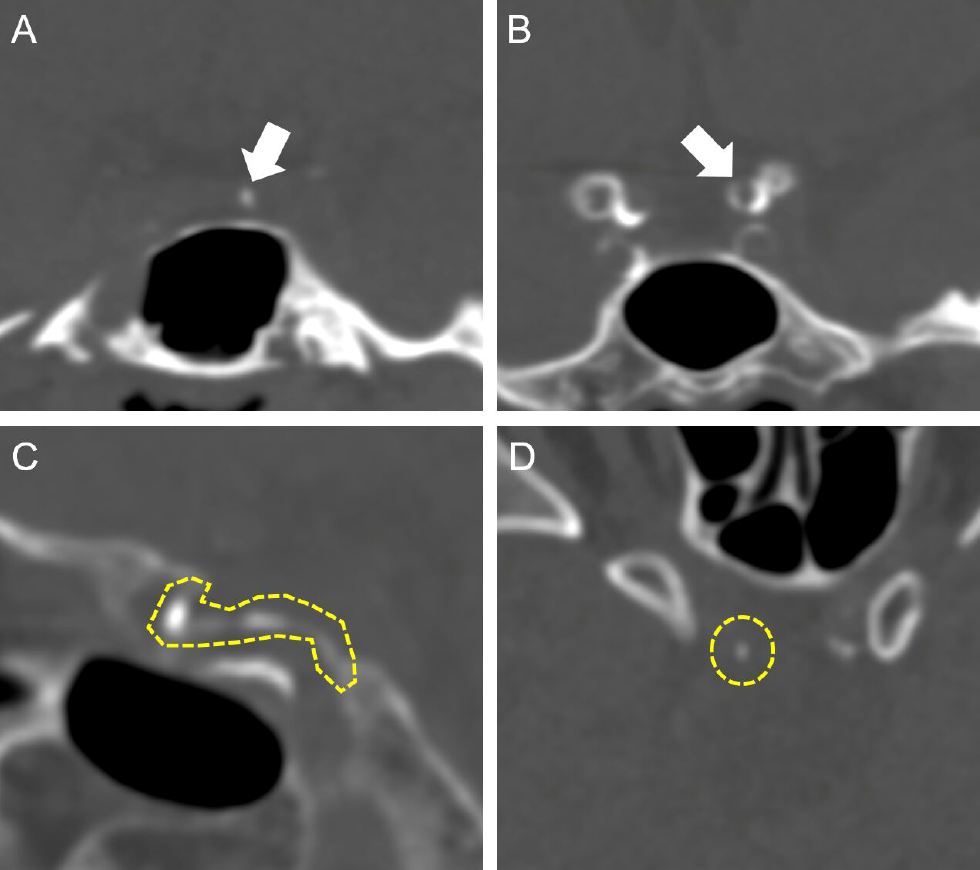
Two patterns of intracranial arterial calcification (IAC) were classified on computed tomography (CT): Intimal IAC (A) and medial IAC (B). The involvement of IAC in each examined artery segment was categorized as diffuse IAC (C) or focal IAC (D).

### 2.3 Evaluation and grading of white matter hyperintensities

The evaluation of WMH was based on T2w and T2 fluid attenuated inversion recovery (T2-FLAIR) sequences. Diffusion weighted imaging (DWI) and apparent diffusion coefficient (ADC) were used to exclude acute infarct lesions. MRI Images were assessed by two independent raters with more than 5 years of experience in brain MRI (H.D. and YJ.D.) who were blinded to clinical characteristics and CT images. White matter hyperintensities (WMH) was graded according to a previously published and validated 8-grade scale^15,16^. Grade 1 referred to discontinuous periventricular rim with minimal dots of subcortical lesions; Grade 2 was thin and continuous periventricular rim with a few patchy subcortical lesions; Grade 3, thicker, continuous periventricular rim with scattered subcortical lesions; Grade 4, thicker, shaggier periventricular rim with mild subcortical lesion (possibly with minimal confluent periventricular lesions); Grade 5 represents for mild periventricular confluent lesion that surrounds the frontal horn and occipital horn; Grade 6 was moderate periventricular confluent lesion surrounding the frontal horn and occipital horn; Grade 7 was periventricular confluent lesion with the centrum semiovale involved moderately; Grade 8, periventricular confluent lesion involving most region of the centrum semiovale. Images with no white matter findings were graded 0 and those with WMH severer than Grade 8 were scored 9. After the grading of WMH, the burden of WMH was categorized as absent, mild, moderate and severe (Figure 2): “WMH absent” indicated Grade 0. “Mild WMH” consisted of Grade 1 and Grade 2. “Moderate WMH”, included Graded 3 to 5. “Severe WMH” comprised of Graded 6 to 9.

**Figure 2.**
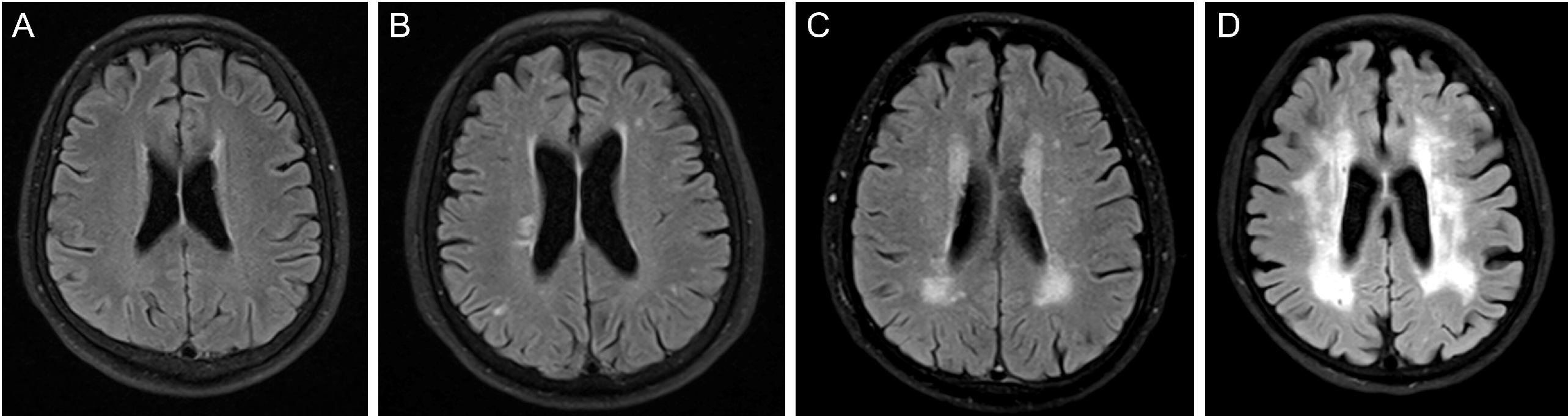
The burden of white matter hyperintensities (WMH): WMH absent (A), mild WMH (B), moderate WMH (C) and severe WMH (D).

### 2.4 Statistical analysis

IBM SPSS (20.0, SPSS, Inc) was used for statistical analysis. Continuous variables were presented as mean ± standard deviation (SD) and categorical variables were presented as numbers and percentages. The inter-rater reliability was assessed by Cohen’s kappa analysis. Independent t-test and Mann–Whitney U test were used for continuous variables and Pearson’s chi-square test and Fisher’s exact test were used for dichotomous categorical variables. Relationships of ordinal variables were measured by Chi-square linear trend test. Vascular risk factors with *p* value < 0.1 were considered as confounding factors and were tested by collinearity diagnostics. Regression analysis was performed before and after adjusting for confounding factors. Relationships between the presence of WMH and presence of IAC (intimal or medial) were examined in multiple binary logistic regression models. Multiple ordinal logistic regression analysis was used to evaluate the relationships of WMH burden with the presence of IAC, the NOA involved by IAC and severity of IAC, respectively. In the ordinal logistic regression models, we set the absence of IAC as reference and the results of regression were interpreted as the risk of larger WMH burden in patients with higher NOA or more severe IAC compared with patients without IAC. A two-sided *p* value < 0.05 was considered statistically significant.

## 3. Results

### 3.1 Baseline characteristics

In total, 265 consecutive stroke patients were included, and 10 patients excluded due to poor image quality or incomplete clinical data. The mean age of the included patients was 62.26 ± 11.99 years and 179 patients were men. Of the 265 patients, 36.2% had history of current smoking and 27.2% had history of drinking. Diabetes was identified in 27.2% of the patients and hypertension was found in 57.0% of the patients. Hyperlipidemia and atrial fibrillation were diagnosed in 16.2% and 4.2% of the patients, respectively. 62 patients had history of previous stroke/TIA and 30 patients had history of ischemic heart disease.

Intracranial arterial calcification was identified in 231 (87.2%) patients. Intimal IAC was present in 169 (54.7%) patients and medial IAC was present in 150 (48.5%) patients. Coexistent (intimal and medial) IAC was found in 52.1% patients. Diffuse IAC was found in 74 patients, of whom all were medial calcifications. WMH was found in 200 (75.5%) patients, among whom 105 patients had mild WMH, 69 patients were categorized in moderate WMH group and 26 patients were identified to have severe WMH. Additional baseline characteristics were presented in Table 1.

**Table 1.** Baseline characteristics of patients with different white matter hyperintensities (WMH) burden.

|  | WMH burden |  |  |  | *p* for trend |
| --- | --- | --- | --- | --- | --- |
|  | Absent (n=65) | Mild (n=105) | Moderate (n=69) | Severe (n=26) |  |
| **Male, n (%)** | 48 (73.8) | 71 (67.6) | 45 (65.2) | 15 (57.7) | 0.125 |
| **Age (mean ± SD, years)** | 51.34 ± 10.60 | 62.64 ± 9.812 | 68.97 ± 9.83 | 70.23 ± 8.16 | <0.001 |
| **Smoking, n (%)** | 31 (47.7) | 45 (42.9) | 28 (40.6) | 8 (30.8) | 0.152 |
| **Drinking, n (%)** | 26 (40.0) | 33 (31.4) | 18 (26.1) | 7 (26.9) | 0.094 |
| **Diabetes, n (%)** | 17 (26.2) | 38 (36.2) | 20 (29.0) | 9 (34.6) | 0.643 |
| **Hypertension, n (%)** | 37 (56.9) | 68 (64.8) | 52 (75.4) | 19 (73.1) | 0.026 |
| **Hyperlipidemia, n (%)** | 18 (27.7) | 16 (15.2) | 12 (17.4) | 4 (15.4) | 0.146 |
| **Atrial fibrillation, n (%)** | 1 (1.5) | 4 (3.8) | 7 (10.1) | 1 (3.8) | 0.107 |
| **Previous stroke/TIA, n (%)** | 6 (9.2) | 23 (21.9) | 24 (34.8) | 9 (34.6) | <0.001 |
| **Ischemic heart disease, n (%)** | 1 (1.5) | 13 (12.4) | 14 (20.3) | 2 (7.7) | 0.026 |

SD, standard deviation. TIA, transient ischemic attack

### 3.2 Association between IAC pattern and WMH

The presence of medial IAC was found to be correlated with the occurrence of WMH (*p*<0.001). Table 2 showed the more relationship after categorizing the burden of WMH. Patients with medial IAC were more prone to have larger WMH burden (*p*<0.001). However, intimal IAC was not correlated with the occurrence of WMH or the burden of WMH.

**Table 2.** Correlations between intracranial arterial calcification (IAC) and white matter hyperintensities (WMH).

|  | Patients,<br>n | WMH occurrence |  |  |  | *p* value |
| --- | --- | --- | --- | --- | --- | --- |
|  |  | Absent, n (%) |  | Present, n (%) |  |  |
| Intimal IAC |  |  |  |  |  |  |
| Absent | 96 | 28 (29.2) |  | 68 (70.8) |  | 0.186 |
| Present | 169 | 37 (21.9) |  | 132 (78.1) |  |  |
| Medial IAC |  |  |  |  |  |  |
| Absent | 115 | 52 (45.2) |  | 63 (54.8) |  | <0.001* |
| Present | 150 | 13 (8.7) |  | 137 (91.3) |  |  |
| WMH burden |  |  |  |  |  |  |
|  |  | Absent, n (%) | Mild, n (%) | Moderate, n (%) | Severe, n (%) | *p* for trend |
| Intimal IAC |  |  |  |  |  |  |
| Absent | 96 | 28 (29.2) | 31 (32.3) | 25 (26.0) | 12 (12.5) | 0.215 |
| Present | 169 | 37 (21.9) | 74 (43.8) | 44 (26.0) | 14 (8.3) |  |
| Medial IAC |  |  |  |  |  |  |
| **Absent** | 115 | 52 (45.2) | 51 (44.3) | 11 (9.6) | 1 (0.9) | <0.001* |
| **Present** | 150 | 13 (8.7) | 54 (36.0) | 58 (38.7) | 25 (16.7) |  |
| **Medial IAC, involved arteries** |  |  |  |  |  |  |
| **0 arteries** | 115 | 52 (45.2) | 51 (44.3) | 11 (9.6) | 1 (0.9) | <0.001* |
| **1 to 3 arteries** | 88 | 12 (13.6) | 33 (37.5) | 31 (35.2) | 12 (13.6) |  |
| **≥ 4 arteries** | 62 | 1 (1.6) | 21 (33.9) | 27 (43.5) | 13 (21.0) |  |
| **Medial IAC, severity** |  |  |  |  |  |  |
| **No medial IAC** | 115 | 52 (45.2) | 51 (44.3) | 11 (9.6) | 1 (0.9) | <0.001* |
| **Focal medial IAC** | 76 | 10 (13.2) | 31 (40.8) | 25 (32.9) | 10 (13.2) |  |
| **Diffuse medial IAC** | 74 | 3 (4.1) | 23 (31.1) | 33 (44.6) | 15 (20.3) |  |

\*: $p < 0.05$

The number of arteries involved by medial IAC was also correlated with the burden of WMH. Patients with larger WMH burden were identified to have higher median number of NOA involved by media IAC (Figure 3). After classifying number of involved arteries into 3 groups according to the quartiles (Table 2), the frequency of larger WMH burden showed a rising trend (Figure 4) among patients with no medial IACs, patients with 1 to 3 arteries involved by medial IAC and patients with at least 4 arteries involved by medial IAC (*p*<0.001). Similar trend was found in the correlations between WMH burden and medial IAC severity (Figure 4). Patients with more severe medial IAC were more likely to have moderate and severe WMH (Table 2), compared with patients with milder medial IAC (*p*<0.001).

**Figure 3.**
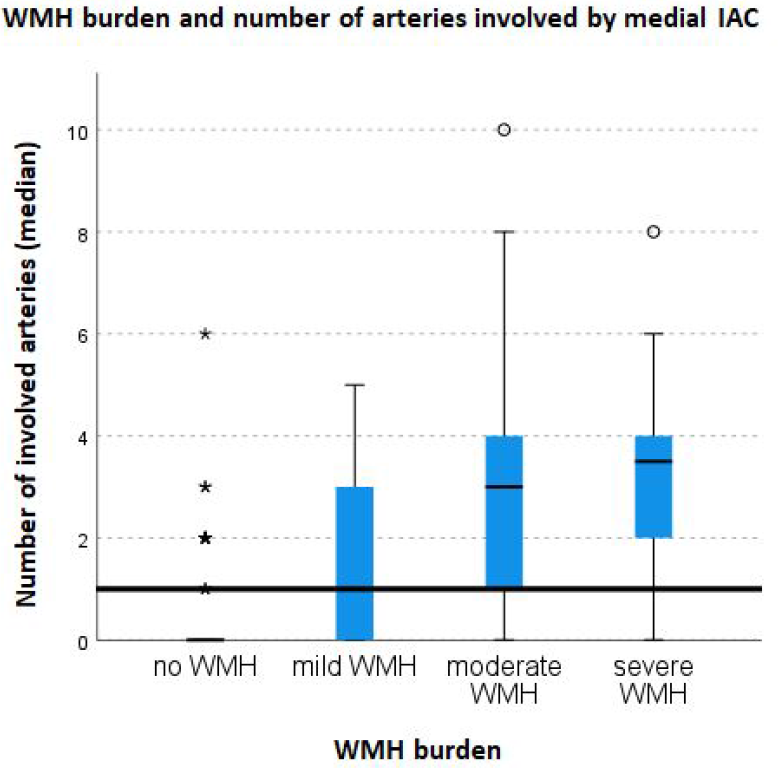
The median number of arteries involved by medial IAC in patients with white matter hyperintensities (WMH). Patients with larger WMH burden were more prone to have more arteries involved by medial IAC.

**Figure 4.**
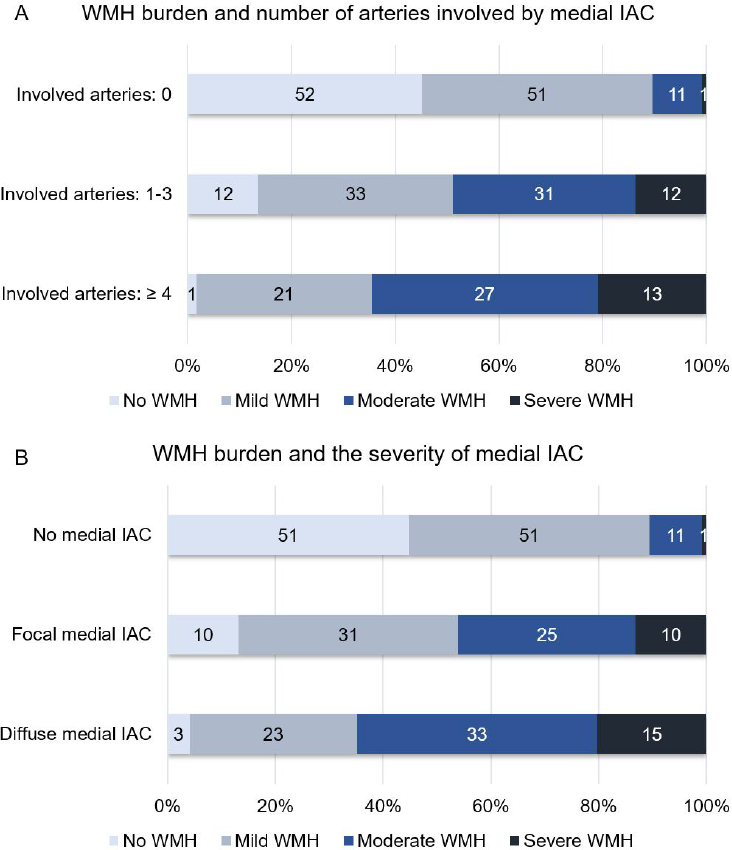
Correlations of white matter hyperintensities (WMH) with the number of arteries involved by medial intracranial arterial calcification (medial IAC) and the severity of medial IAC. Patients with medial IAC embedded in more artery segments (A) and patients with more severe medial IAC (B) tended to have larger WMH burden.

Binary logistic regression suggested that the presence of medial IAC was correlated with the occurrence of WMH (crude OR, 8.698; 95% CI, 4.420-17.119; *p*<0.001). After adjusting for confounding factors (age, hypertension, previous stroke/TIA and ischemic heart disease), multiple binary logistic regression indicated that the correlation remained statistically significant (adjusted OR, 3.794; 95% CI, 1.741-8.267; *p*<0.001).

The relationships of WMH burden with the presence of medial IAC, the NOA involved by medial IAC and the severity of medial IAC were examined by ordinal logistic regression models (Table 3). The presence of medial IAC suggested higher risk of larger WMH burden, before (crude OR, 9.839; 95% CI, 5.719-16.629; *p*<0.001) and after (adjusted OR, 4.947; 95% CI, 2.780-8.801; *p*<0.001) adjusting for confounding factors. Compared with the absence of medial IAC, 1 to 3 arteries involved by medial IAC (adjusted OR, 3.956; 95% CI, 2.141-7.310; *p*<0.001) and 4 or more arteries involved by medial IAC (adjusted OR, 7.215; 95% CI, 3.549-14.667; *p*<0.001) indicated higher risk of larger WMH burden, after adjusting for confounding factors. Similarly, compared with the absence of medial IAC, focal medial IAC (adjusted OR, 3.966; 95% CI, 2.107-7.466; *p*<0.001) and diffuse medial IAC (adjusted OR, 6.506; 95% CI, 3.297-12.841; *p*<0.001) also correlated with higher risk of larger WMH burden, after adjusting for confounders.

**Table 3.**
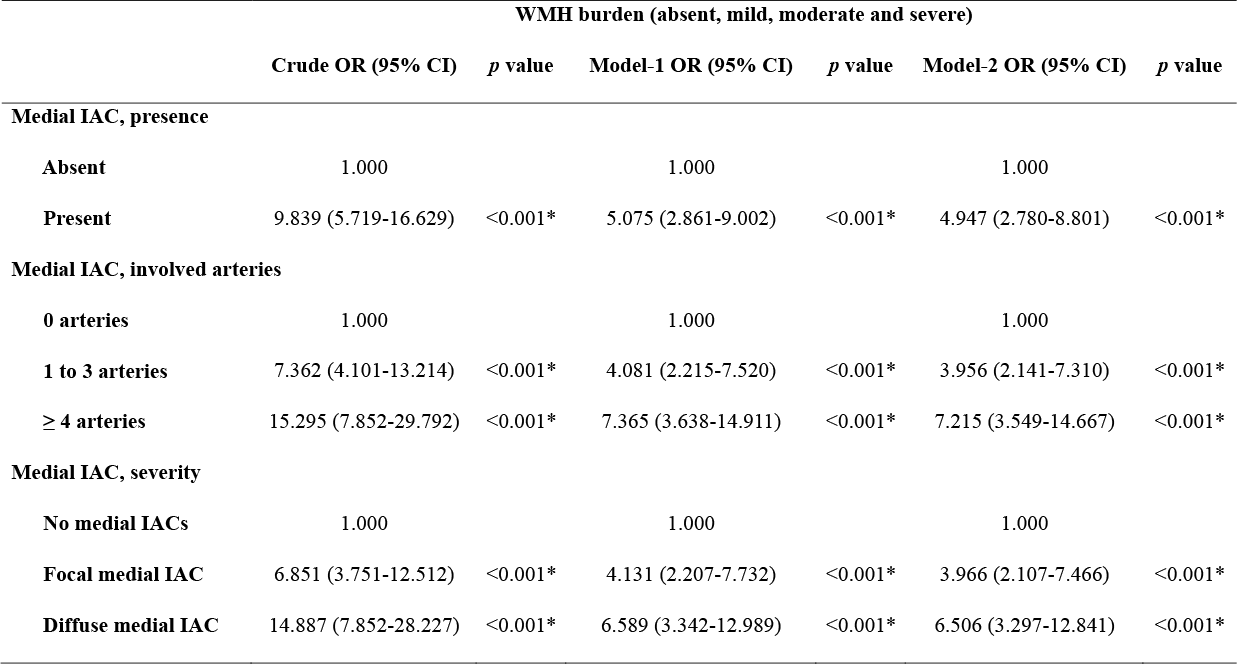
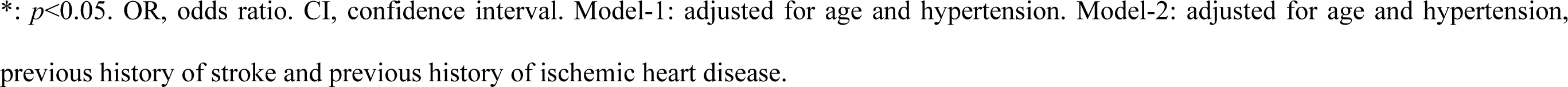
Multiple ordinal logistic regressions on the correlations between medial intracranial arterial calcification (IAC) and white matter hyperintensities (WMH) burden.

|  | WMH burden (absent, mild, moderate and severe) |  |  |  |  |  |
| --- | --- | --- | --- | --- | --- | --- |
|  | Crude OR (95% CI) | *p* value | Model-1 OR (95% CI) | *p* value | Model-2 OR (95% CI) | *p* value |
| **Medial IAC, presence** |  |  |  |  |  |  |
| **Absent** | 1.000 |  | 1.000 |  | 1.000 |  |
| **Present** | 9.839 (5.719-16.629) | <0.001* | 5.075 (2.861-9.002) | <0.001* | 4.947 (2.780-8.801) | <0.001* |
| **Medial IAC, involved arteries** |  |  |  |  |  |  |
| **0 arteries** | 1.000 |  | 1.000 |  | 1.000 |  |
| **1 to 3 arteries** | 7.362 (4.101-13.214) | <0.001* | 4.081 (2.215-7.520) | <0.001* | 3.956 (2.141-7.310) | <0.001* |
| **≥ 4 arteries** | 15.295 (7.852-29.792) | <0.001* | 7.365 (3.638-14.911) | <0.001* | 7.215 (3.549-14.667) | <0.001* |
| **Medial IAC, severity** |  |  |  |  |  |  |
| **No medial IACs** | 1.000 |  | 1.000 |  | 1.000 |  |
| **Focal medial IAC** | 6.851 (3.751-12.512) | <0.001* | 4.131 (2.207-7.732) | <0.001* | 3.966 (2.107-7.466) | <0.001* |
| **Diffuse medial IAC** | 14.887 (7.852-28.227) | <0.001* | 6.589 (3.342-12.989) | <0.001* | 6.506 (3.297-12.841) | <0.001* |

\*: $p < 0.05$ . OR, odds ratio. CI, confidence interval. Model-1: adjusted for age and hypertension. Model-2: adjusted for age and hypertension, previous history of stroke and previous history of ischemic heart disease.

### 3.3 Interrater reliability of assessments on IAC and WMH

The inter-rater reliabilities of IAC presence, IAC pattern (intimal and medial) and IAC severity (focal or diffuse) were evaluated separately. The weighted kappa of IAC presence was 0.904 (95% CI, 0.839-0.969, *p*<0.001), the weighted kappa of IAC pattern was 0.877 (95% CI, 0.760-0.993, *p*<0.001) and the weighted kappa of IAC severity was 0.871 (95% CI, 0.747-0.995, *p*<0.001). WMH occurrence and WMH burden were also evaluated separately. The weighted kappa of WMH occurrence was 0.932 (95% CI, 0.802-1.063, *p*<0.001) and the weighted kappa of WMH burden was 0.860 (95% CI, 0.730-0.990, *p*<0.001).

## 4. Discussion

In summary, this study for the first time identified that the number of intracranial arteries involved by medial IAC and the severity of medial IAC were independently associated with the presence and burden of WMH while intimal IAC did not correlate to WMH. With the number of involved arteries and severity of medial IAC being higher, the burden of WMH was heavier. These findings may suggest that medial pattern of calcification in intracranial vascular beds may be a potential biomarker for CSVD.

The inherent difference between intimal IAC and medial IAC may be correlated with different vascular consequences. Intimal IAC is associated with large artery atherosclerosis^11,12^ while medial IAC is more related to arterial stiffness^13,17^, which may cause impaired vasodilation and vessel compliance. Although several studies suggested possible correlations between intracranial atherosclerosis and WMH^18-20^, in consideration of distinct etiology of large artery atherosclerosis and CSVD, such correlations are more likely explained by mutual risk factors such as hypertension and aging^21,22^. CSVDs are mainly systemic vascular disorders and may occur without hypertension^23^, suggesting more complex pathogenesis than arteriolar occlusions. In ischemia led by CSVD, restricted vessels may lead to a state of chronic hypoperfusion in the microvascular beds that supplies the white matter. Eventually, the myelinated fibers degenerate as a result of repeated oligodendrocyte damage^24^. One of the causative roles in white matter lesions may be altered intracranial vascular pulsatility^25^, which could result from the progressing arterial stiffness. As medial IAC evolves widely in multiple intracranial vessel beds, the degree of global arterial stiffness may advance, thus leading to restricted vasodilation and hypoperfusion in vast subcortical regions during needs of increased neuronal activity and metabolic or vasodilatory challenge. On the other hand, increased pulsatile wave transmission from the aorta to the microcirculation caused by arterial stiffness may also be hazardous to cerebral microvasculature^26^. In line with our findings, a recent study identified that the volume of medial IAC, rather than that of intimal IAC, mediates the association between blood pressure and WMH^27^. These findings indicate that medial IAC, as a proxy for arterial stiffness, may play an independent role in the pathogenesis of CSVD. Given the fact that evaluation of subcortical cerebrovascular activity is difficult, our results may benefit the understanding of CSVD from a different aspect.

Associations between IAC and cognitive disorders were reported. Larger IAC volume was found to be correlated with worse cognitive performance^28^ and higher risk of cognitive decline^29^. Cognitive impairment was found to be associated with IAC in patients with chronic kidney disease that may cause irregular serum calcium^30^. However, despite the high prevalence of IAC in patients with cognitive disorders, IAC may not be directly related to cognitive outcome^31,32^. The bridge that connects IAC with cognitive impairment could be CSVD. White matter lesions led by CSVD have been found to be a predictor of cognitive decline^33^ and dementia^34^. By far, neuroimaging has revealed notable association between WMH and cognitive impairment^35,36^. The total WMH volume is a strong predictor of cognitive impairment^37^. Increasing WMH burden is correlated with diffuse loss of cortex and white matter in vast brain areas, resulting in reduced global network efficiency^23^ and worse cognitive performance^38^. Moreover, WMH also have impact on movement disorder^39^ and neuropsychiatric symptoms^40^ in people with cognitive disorders. By identifying medial IAC as a potential biomarker for CSVD, this study could provide new perspectives about the correlation between IAC and cognitive impairment in future studies.

Identifying medial IAC as an imaging biomarker of CSVD may also contribute to the determination of potential candidates for additional MRI scanning among admitted patients. CSVD accounts for nearly half of all dementias^23^, of which the economic burden is high. In China, the annual total national cost of dementia ranges from $ 69 billion to $ 195 billion^41,42^ and the expense per patient tends to rise with the progression of dementia^43^. Considering population aging and the increasing prevalence of dementia, early detection of CSVD and CSVD-related dementia is essential for reducing costs. The symptoms and clinical course of CSVD are highly variable^23^, hence medial IAC, as a potential imaging biomarker, may be helpful during patient screening. In real-world clinical activities, medical supplies, especially imaging assessments, are invariably inadequate to fully cover every aspect for individuals. As a result, studies on the cost-effectiveness of brain MRI modality for patients with potential CSVD may be needed in the future. It has been reported that in patients with minor stroke, additional brain MRI scanning has been found to enable timely secondary prophylaxis, which results in lower costs and higher cumulative quality-adjusted life years^44^. In light of the heavy burden of CSVD-related functional disability and cognitive impairment, appropriate brain MRI scanning will most likely ensure early detection and timely interventions on CSVD, thereby reducing long-term expense and improving final outcomes.

There are several limitations in this present study. First, this is a single-center study, which could lead to potential selection bias. Second, all patients included in this study had acute ischemic stroke. Therefore, there might be an overestimated rate of IAC and WMH. Third, as a cross-sectional study, the identified association may lack the capacity to infer causative roles of medial IAC on the pathogenesis of WMH. Future studies may be needed.

## 5. Conclusions

Our findings on this cohort of acute stroke patients demonstrated that medial IAC was correlated with the occurrence of WMH. The number of arteries involved by medial IAC and the severity of medial IAC are correlated with the burden of WMH, suggesting dose-effect relationships between medial IAC and WMH burden. These findings indicate a potential impact of artery calcification on the pathogenesis of cerebral small vessel disease. Further studies are needed to investigate the shared underlying mechanisms of intracranial large artery disease and cerebral small vessel disease.

## Data Availability

The data referred to in the manuscript is available.

## 6. Author contribution

All authors equally contributed to the conception and design of this study. Material preparation, data collection and data preservation were performed by Jianrong Zheng, Cong Liu, Yajing Cheng, and Jun Hu. Data analysis was performed by Heng Du, Xuelong Li and Yanjing Dong. The first draft of this manuscript was written by Heng Du. Manuscript revisions were conducted by Heng Du under the advice of Jun Hu and Xiangyan Chen. All authors have approved the submission of this manuscript.

## 7. Disclosures

The authors declare no conflict of interest in this study.

## Notes

### Competing Interest Statement

The authors have declared no competing interest.

### Funding Statement

The present study is not supported by any funding.

### Author Declarations

This study was approved by the Clinical Research Ethics Committee of the Peking University Shenzhen Hospital and the Ethics Committee of The Hong Kong Polytechnic University.

